# Hierarchical Graph Learning with Multimodal Distillation for Breast Cancer Survival Prediction

**DOI:** 10.64898/2026.11.14.26368121

**Authors:** Binh Minh Tran, Van An Nguyen, Quynh Anh Dang, Minh Hai Nguyen Ngoc, Tran Hai Nam, Dang Hung Tran

## Abstract

Imaging mass cytometry (IMC) and DNA methylation can provide complementary information for breast cancer prognosis, yet these measurements may be unavailable for new patients. In this paper, we propose HEAL-Surv, a teacher-student framework that uses both modalities during training to predict five-year net disease-specific survival (DSS) from clinical variables and molecular profiles at inference. An additive Cox teacher integrates all four modalities, using a hierarchical IMC graph encoder to capture local cell relationships and coarser tissue organisation. The student learns from observed outcomes and cross-fitted teacher survival curves, giving more weight to targets that agree across random initialisations. In five-fold cross-validation on 423 METABRIC patients, the full teacher achieved a mean five-year Uno’s concordance of 0.723. Removing any modality reduced mean concordance and increased mean Brier-based errors. Relative to the matched student trained on observed outcomes alone, distillation increased mean five-year Uno’s concordance from 0.658 to 0.704, with higher integrated and five-year Brier scores. In a separate external evaluation, an IMC-clinical variant trained on METABRIC achieved a five-year Uno’s concordance of 0.660 on 265 Basel patients without refitting. These findings support multimodal supervision for improving risk discrimination with fewer inference inputs, while external validation of the student remains necessary.

## 1 Introduction

Breast cancer survival prediction aims to estimate the probability that a patient survives beyond a given time. Patients with similar clinical characteristics can experience different outcomes, motivating the use of molecular profiles alongside clinical variables [11,25]. The computational task is to learn a patient-specific survival curve from these measurements and outcome data that include right-censored observations. When follow-up ends without a recorded breast cancer death, the patient’s observed survival time still informs the model.

Cox proportional-hazards (CPH) models [10], random survival forests (RSF) [19], and neural survival models such as DeepSurv [22] and DeepHit [23] typically represent each patient as a feature vector. This representation readily accommodates clinical variables and molecular profiles. When tissue measurements are aggregated into a single vector, however, information about which cells are neighbours and how those neighbourhoods are organised can be lost. Learning from tissue architecture therefore requires a representation that preserves spatial relationships.

Imaging mass cytometry (IMC) measures protein markers at single-cell resolution while retaining cell locations within tissue [15]. Associations between cellular organisation and clinical outcomes motivate the use of IMC in survival modelling [1,12,20]. Spatial graphs provide a way to retain this organisation by representing cells as nodes and their spatial relationships as edges. DMGN combines regional IMC embeddings with embeddings of patient-level variables [14], and SpaCEy learns cell graphs from protein features [26]. CytoCommunity [17] identifies cellular neighbourhoods, and HRCHY-CytoCommunity [32] extends this approach to hierarchical tissue organisation. These studies motivate an IMC representation that captures both local cell relationships and the organisation of neighbourhoods across tissue regions.

Tissue architecture is one source of information that a survival model can combine with other patient measurements. Pathomic Fusion [8], MCAT [9], and SurvPath [21] integrate histological and molecular information through feature fusion, genomics-guided attention, and pathway-tissue interactions, respectively. DNA methylation provides a further view of breast cancer heterogeneity [3]. The measurements available for model development may be more extensive than those available for a new patient. We study a setting in which clinical variables and molecular profiles are available at inference, with IMC and DNA methylation available during training.

Knowledge distillation accommodates this difference in input availability by allowing a teacher to use additional modalities when supervising a student. Previous studies have used privileged modalities in medical imaging [7] and transferred transcriptomic information to pathology-based survival models [33]. We ask whether supervision from a teacher with access to IMC and methylation improves a student that already uses clinical and molecular inputs. We compare students with the same architecture and inference inputs, one trained on observed outcomes alone and one also trained on teacher predictions.

In this paper, we propose HEAL-Surv, a teacher-student framework for five-year net DSS prediction. The teacher combines a hierarchical IMC graph representation with clinical variables, molecular profiles, and methylation features in an additive Cox model. The molecular-clinical student learns from observed outcomes and cross-fitted teacher survival curves, with target weights based on agreement across random initialisations. We evaluate whether distillation improves risk discrimination and survival-probability accuracy.

1. We combine edge-aware GATv2 [6] with two-stage MinCutPool [4] to encode local cell relationships and coarser tissue organisation. The additive Cox teacher integrates this IMC representation with molecular, methylation, and clinical features.
2. We train a molecular-clinical student on observed outcomes and cross-fitted teacher survival probabilities at years 1-5. Each patient’s targets come from teachers fitted and selected on the other folds and receive weights based on agreement across random initialisations. The student predicts from molecular and clinical inputs alone.
3. In 423 METABRIC patients, we assess risk discrimination and survival-probability error through matched student comparisons and teacher ablations of modalities and hierarchical encoding. In 265 Basel patients, we externally validate the IMC-clinical variant and examine its spatial behaviour.

## 2 Methodology

HEAL-Surv has two training stages. We first fit an additive Cox teacher to observed outcomes using hierarchical IMC graph representations together with molecular, methy-lation, and clinical features. We then fit a molecular-clinical student using observed outcomes and cross-fitted teacher survival curves, with greater weight given to targets that agree across initialisations.

### 2.1 Problem Formulation and Framework Overview

For patient *i*, **g**_*i*_ contains expression, mutation, and copy-number alteration (CNA) features. The vectors **m**_*i*_ and **c**_*i*_ contain methylation and clinical features, respectively. We represent IMC measurements as 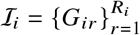 , with one cell graph for each tissue core. Oestrogen receptor (ER) status defines stratum *s*_*i*_ ∈{ER+ , ER™} and is recorded separately from **c**_*i*_. The dataset is

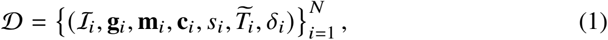

where the observed outcome is 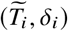. Let *T*_*i*_ denote the latent time from diagnosis to breast cancer death and *C*_*i*_ the censoring time, determined by other-cause death or last follow-up. Then 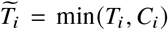 and *δ*_*i*_ = I (*T*_*i*_ ≤ *C*_*i*_) , so *δ*_*i*_ = 0 indicates a right-censored observation. For patient inputs *X* _*i*_, the prediction target is the conditional survival function *S* (*t*| *X* _*i*_ = Pr *T*_*i*_ > *t*|*X* _*i*_) over 0 ≤*t* ≤*τ* for the primary prediction task, with *τ* = 5 years. We estimate this function from event times and right-censored observations. The teacher and student use separate mappings for their respective inputs.

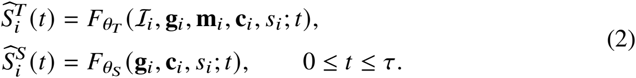

Here *θ*_*T*_ and *θ*_*S*_ are separate model parameters. The student is trained using both observed outcomes and teacher predictions, so IMC and methylation contribute super-vision without entering its prediction mapping. We assume conditionally independent censoring, *T*_*i*_ ⊥ *C*_*i*_ | *X*_*i*_. With other-cause deaths treated as censored observations, the curves represent net DSS rather than cumulative breast cancer mortality under competing risks [2].

Figure 1 shows how these mappings are implemented. First, each IMC core is converted into a cell graph (panel c). Nodes describe protein profiles and cell phenotypes; edges connect spatial neighbours and encode their pairwise attributes. The hierarchical encoder (panel d) applies edge-aware GATv2 message passing, followed by two Min-CutPool stages that coarsen the graph to 12 and then 3 groups, with further message passing at each coarse level. Readouts from the cellular and coarse levels, together with tissue-composition summaries, form a core embedding. Cell-count-weighted aggregation across cores then gives one IMC embedding per patient.

**Fig 1.**
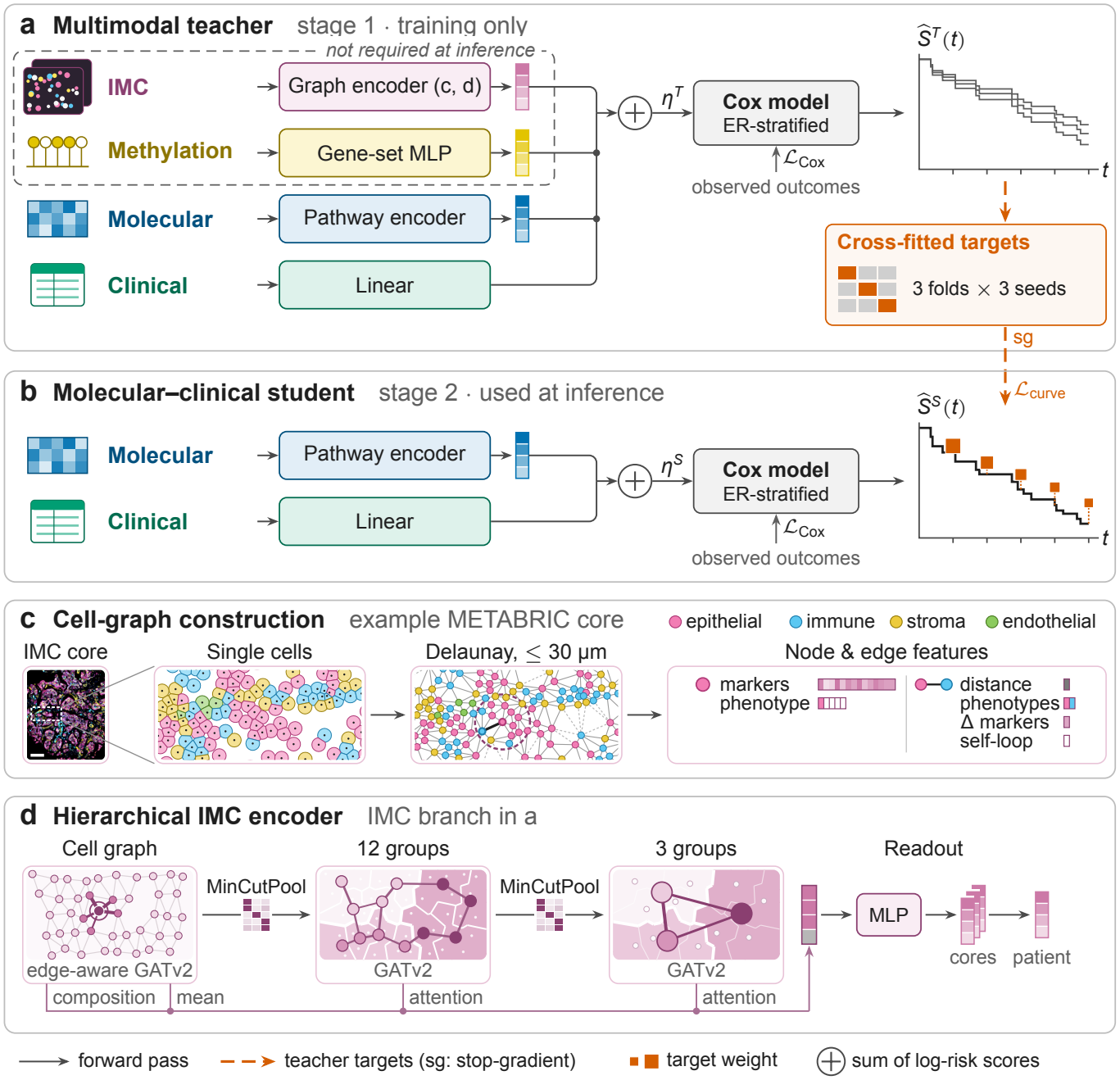
HEAL-Surv: (a) teacher; (b) student; (c) IMC cell graph (MB0282_1 [12]); (d) hierarchical encoder.

In parallel, the molecular branch encodes pathway-level expression and genomic alterations, the methylation branch encodes gene-set summaries, and the clinical branch computes a linear score (panel a). The teacher projects each nonclinical embedding to a scalar log-risk score and sums the four branch scores to obtain 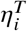. An ER-stratified Cox model combines this score with the estimated ER-specific cumulative baseline hazard to produce the teacher survival curve 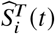.

We generate student targets by three-fold cross-fitting with three teacher initialisations per fold complement. For each patient, the mean survival probabilities at years 1-5 come from teachers fitted and selected without that patient. The student combines the Cox loss on observed outcomes, ℒ_Cox_, with a curve-matching loss, ℒ_curve_, that weights eligible targets by agreement across initialisations (panel b).

At inference, only the student’s molecular and clinical branches are evaluated. Their summed log-risk score 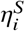and the patient’s ER status are combined with the student’s own frozen ER-specific baseline to output 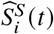, including the five-year net DSS probability 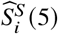. Neither IMC, methylation nor the teacher is required for this step. The following subsections detail the encoders, teacher and distillation objectives.

### 2.2 Edge-Aware Hierarchical IMC Encoder

Core graphs connect Delaunay neighbours within *r*_0_ = 30 *µ*m using calibrated cell centroids. Nodes combine protein vectors **x**_*u*_ ∈ℝ^*P*^ and one-hot cell types **y**_*u*_. Markers are clipped at fitting-set 99.9th percentiles, arcsinh-transformed with cofactor one, and standardised. Omitting patient/core indices, each directed edge *v* →*u* carries

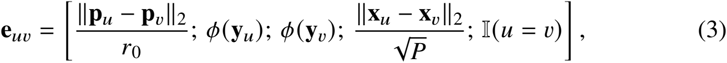

where **p**_*u*_ denotes the cell centroid and *ϕ* maps each cell type to a learned four-dimensional embedding. The 11 attributes encode distance, ordered cell types, protein difference, and self-loop status. We include both directions for each spatial edge. Self-loops have zero distance and protein difference and are excluded from the pooling adjacency.

Edge-aware GATv2 [6] conditions aggregation on these relationships. Starting from **h**_*u*_ = [**x**_*u*_; **y**_*u*_], it computes

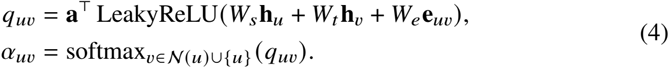

*N* (*u*) denotes spatial neighbours; *W*_*s*_, *W*_*t*_ , *W*_*e*_, **a** are learned. Each head aggregates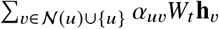head outputs are concatenated. Two four-head layers with ELU,dropout 0.1, projected residuals and layer normalisation yield 64-dimensional cell states*H* ^(0)^.

MinCutPool [4] summarises coarser structure in two stages: *K*_1_ = 12 groups,then *K*_2_ = 3. For zero-diagonal adjacency *A*, initialise 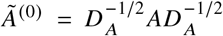 with *D* _*A*_ = diag(*A***1**) and zero inverse degrees for isolated nodes. At level *j* ∈ {0, 1},

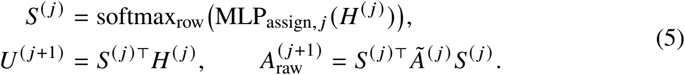

With *D* ^(*j*)^ = diag(*Ã*^(*j*)^ **1**), assignment regularisation is

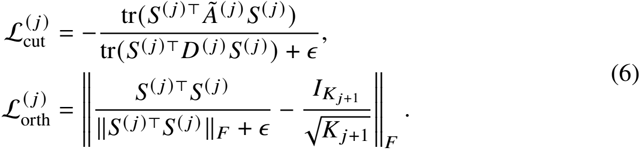

These terms encourage within-group connectivity and nondegenerate assignments. We use *ϵ* = 10^™8^ and zero cut loss for edgeless levels. Removing pooled diagonals and symmetrically normalising yields *Ã*^(*j*+1)^.

Group sizes propagate as soft counts: **n**^(0)^ = **1** and **n**^(*j*+1)^ = *S* ^(*j*)⊤^**n**^(*j*)^. Concatenated LN (*U* ^(*j*+1)^)and(log 1 +**n**^(*j*+1)^)are projected and processed by one coarse GATv2 layer, yielding *H* ^(*j*+1)^. Coarse edge attributes are normalised adjacency weights and self-loop indicators. Groups need not be anatomically connected or biologically equivalent across cores.

Attention pooling [18] summarises each coarse level:

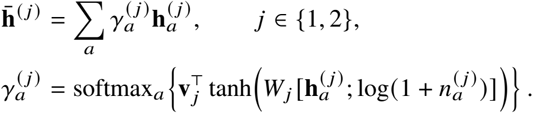

Core readout combines cellular, coarse-level and composition summaries; patient readout weights cores by cell count:

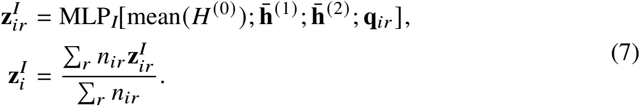

*n*_*ir*_ counts cells; **q**_*ir*_ contains cell-type proportions and mean processed marker levels.

### 2.3 Molecular, Methylation and Clinical Encoders

The molecular branch combines pathway-level expression with genomic alterations. Log-scale expression is mapped from probes to genes using fixed annotations, filtered and standardised. Hallmark sets [24] restrict pathway units to retained member genes *G*_*p*_:

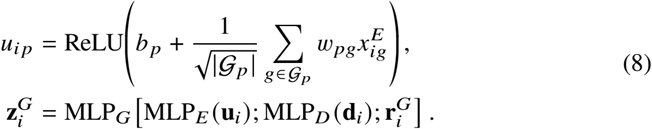

Here 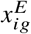is standardised expression; **d**_*i*_ contains mutation indicators and signed CNA; and 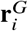 flags missingness, including imputed mutation calls. Empty pathways are omitted and non-member connections fixed to zero. Teacher and student use separate parameters.

After filtering promoter measurements by coverage, we median-impute missing values, standardise the values, and average them within prespecified gene sets. An MLP maps these summaries and the fractions of retained promoter measurements missing before imputation to 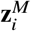. The primary cohort includes patients with an available methylation assay.

In the primary METABRIC analysis, the linear score 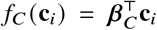 uses age standardised on the fitting set and binary indicators for chemotherapy (CT), endocrine therapy (HT), and radiotherapy (RT). All four covariates and ER status are required. ER selects the baseline stratum, and the student fits its own clinical coefficients. We use retrospective treatment records as prognostic covariates. Use at diagnosis would require verifying their availability at that time. Estimating treatment effects would require a separate study design.

### 2.4 Additive Multimodal Survival Teacher

Adding linear projections of the three embeddings to the clinical score defines an ER-stratified Cox model [10]:

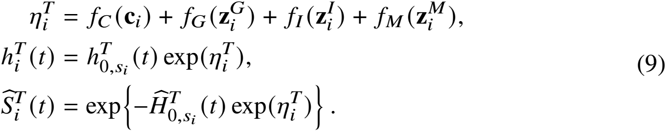

ER strata share score parameters but have separate baseline hazards. Proportional hazards are assumed within strata. Additive fusion excludes IMC-molecular feature exchange. Joint training minimises

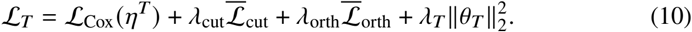

Here ℒ_Cox_ is the negative partial log-likelihood per event, evaluated on complete within-stratum risk sets with Efron ties [13]. Pooling losses are averaged over levels,cores per patient and patients.

Each model estimates its baseline from fitting patients P using Breslow’s estimator [5]:

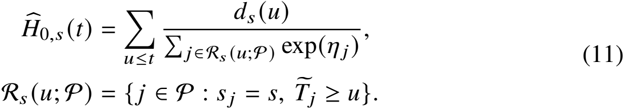

Here *u* denotes an event time, *d*_*s*_ *u* counts events at that time in stratum *s*, and *η* _*j*_ is patient *j* ’s log-risk score. Each fitting stratum must contain at least one event. We freeze the estimated cumulative baselines for evaluation and use the appropriate baseline to convert each patient’s score to a net risk estimate comparable across strata.

### 2.5 Survival-Aware Knowledge Distillation

We divide the student fitting set ℘ into three folds. For each held-out fold ℘_*k*_, we train three independently initialised teachers on ℘_™*k*_ = ℘ \ ℘_*k*_ and predict survival curves for patients in ℘_*k*_. Teacher preprocessing, model selection, stopping, and baseline estimation use only℘_*− k*_. At *T* = {*t*_1_, … , *t*_*L*_} = {1, … , 5}years, we compute the mean and variance across the three predictions for each patient.

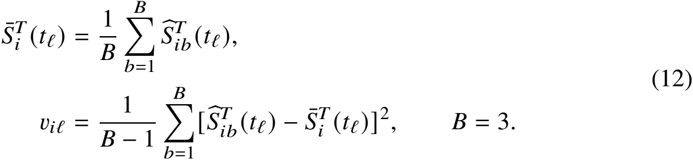

We set *a*_*il*_ = 1 if at least ten patients in stratum *s*_*i*_ remain at risk at *t*_*l*_ and the estimated censoring survival at that time exceeds 0.05, and set it to zero otherwise. Both checks use only the fold complement. We use between-seed variance as a measure of initialisation stability. Calibrated predictive uncertainty would require a separate assessment.

The student uses molecular and clinical scores with its own baseline:

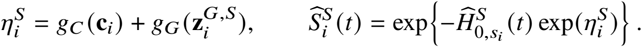

We assign eligible targets weights *w*_*il*_ = max {0.1, (1+ *v*_*il*_ /*v*_0_) ^™1^}. Weights decrease with variance across initialisations until they reach the floor of 0.1. We set *v*_0_ to the median positive variance among eligible targets in ℘ , with a lower bound of 10^™6^. If all eligible variances are zero, we assign unit weights. The curve-matching loss is

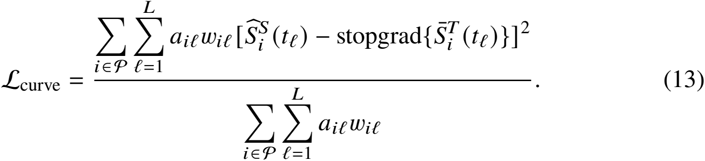

We keep the teacher targets, masks, and weights fixed during student training. The curve-matching loss is zero when no patient-time pair satisfies the support criteria. A teacher target may extend beyond a patient’s observed follow-up. The Cox term uses the observed event or censoring outcome, and the curve term uses the eligible teacher predictions. The student objective is

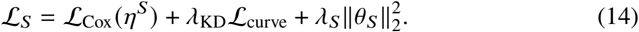

At each full-batch update, we recompute the student’s cumulative Breslow baseline from its current scores and the full fitting-set risk sets, differentiating through the scores and baseline denominators. After fitting, we recompute and freeze the baseline for evaluation. We set *λ*_KD_ = 0 for the matched student without distillation and assign unit weights to eligible targets for the uniform-distillation student. We use observed validation outcomes for model selection (Algorithm 1).

## 3 Experimental Setup

### 3.1 Cohorts and Evaluation Protocol

#### Algorithm 1

HEAL-Surv within one outer fold

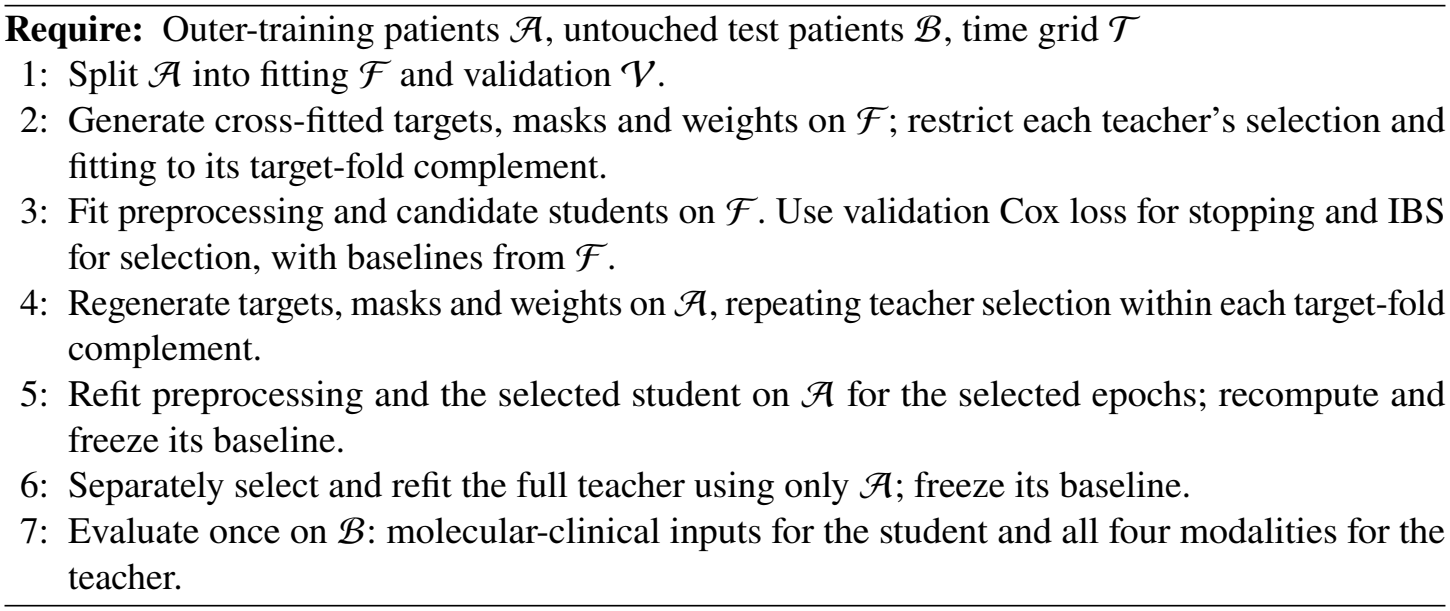

The matched METABRIC cohort links clinical, molecular, IMC and promoter-methylation records for 423 patients through released identifiers [1,3,11,25]. The Basel cohort comprises 265 patients with clinical and IMC records [20] (Table 1). Within each cohort, we join records by exact matches on normalised patient or sample identifiers, verify sample groupings by patient, and reject duplicate or conflicting identifiers. We retain all eligible tissue cores and keep each patient’s cores and assays in the same partition. Feature selection, preprocessing, and imputation use fitting patients only. Both cohorts use the DSS definition in Sect. 2.1, with follow-up measured in years and a primary horizon of five years. The primary METABRIC comparisons include patients with all four modalities, valid follow-up, and complete clinical covariates.

**Table 1.** METABRIC and Basel cohort characteristics.

| **Characteristic** | **METABRIC** | **Basel** |
|---|---|---|
| Patients analysed, *n* | 423 | 265 |
| Survival endpoint | DSS | DSS |
| Outcome events, *n* | 138 | 57 |
| Events within 5 years, *n* | 73 | 40 |
| Patients at risk at 5 years, *n* | 305 | 175 |
| Prediction horizon, years | 5 | 5 |
| Median follow-up, years | 12.33 | 7.92 |
| Age, median (IQR), years | 60.3 (51.6-70.1) | 62.0 (51.0-73.0) |
| ER-positive, *n* (%) | 343 (81.1) | 206 (77.7) |
| IMC tissue images, *n* | 467 | 273 |
| Clinical variables | Age, ER, CT, HT, RT | Age, ER, CT, HT |
| Available modalities | Clinical, molecular, IMC, methylation | Clinical, IMC |

For the primary METABRIC analysis, IMC uses 36 protein markers and 32 cell-type categories. Its markers are clipped at fitting-set 99.9th percentiles (Sect. 2.2). The four clinical covariates are age, CT, HT and RT; ER defines the baseline stratum. Across the five outer METABRIC fitting sets, retained feature counts are 20,192 for expression,22,544 for CNA, 152-158 for mutation and 20,705-21,025 promoter rows for methylation. For non-IMC modalities, entirely missing features are removed, remaining values are median-imputed, and zero-variance features are discarded. Methylation measurements require coverage ≥1.

METABRIC uses five outer patient-level folds, an 80/20 fitting/validation split within outer training and three-fold teacher cross-fitting (Algorithm 1). Splits are jointly stratified by ER and observed-event status at every level. Matched students share architecture and inference inputs; teacher modality ablations use the same patients and folds. The clinical ablation removes both covariates and ER. We repeat five-fold cross-validation three times in Basel. The clinical reference models use age, ER, CT, and HT. DMGN and the HEAL-Surv spatial variant also use IMC, and the additional CPH reference combines IMC-derived protein features with the clinical inputs. Cox-based models use ER-specific baselines, while the other references include ER as a binary predictor.

The component ablation protocol uses the same 423 METABRIC patients, five outer folds, fitting/validation partitions and training-seed schedule. Uniform KD retains the student architecture, cross-fitted teacher targets and support masks, setting *w*_*il*_ = 1 for supported targets. Both KD variants use identical search spaces and selection procedures, independently selecting *λ*_KD_ within training data.

The non-hierarchical teacher retains the cell graphs, cell-level edge-aware GATv2, composition summaries and other modality branches. It replaces the two-stage MinCut-Pool branch with direct core readout:

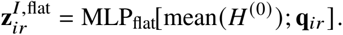

Both teacher variants use the same IMC output dimension, cell-count-weighted patient aggregation, and tuning budget. The flat variant omits the coarse GATv2 layers and pooling penalties, so the comparison evaluates the hierarchical branch as a whole.

The external evaluation uses age, CT, HT, and ER status, which are available in both cohorts. ER defines the Cox strata or enters the other models as a binary predictor. The IMC-based models use six markers matched by target and clone, with cell types mapped to five broad phenotype groups. In Basel, CT records chemotherapy before or after surgery, and HT records postoperative endocrine therapy. We fit preprocessing on METABRIC fitting patients and freeze it along with the model parameters and Cox baselines before applying the IMC-clinical variant and shared-input references to Basel.

### 3.2 Metrics

Uno’s five-year concordance [29] is the primary discrimination measure, and Harrell’s concordance is secondary. For both measures, we rank patients by their predicted five-year net risk, 1 − Ŝ_*i*_ 5 , using ER-specific baselines where applicable. We measure probability error using the five-year Brier score and the integrated Brier score (IBS) over a monthly grid from 1 to 60 months, with inverse probability of censoring weights (IPCW) [16]. Higher concordance indicates better risk ranking, and lower Brier scores indicate smaller probability errors. Brier scores reflect both discrimination and calibration [30].

For within-cohort evaluation, we report means and standard deviations (SDs) across five METABRIC test folds and 15 Basel test folds. For external evaluation, we report SDs across 1,000 paired patient-level bootstrap samples of the fixed predictions, keeping each patient’s cores together. We estimate censoring in the external cohort solely to compute the metrics. The fold and bootstrap SDs describe different sources of variability, and we interpret the comparisons descriptively.

### 3.3 Implementation Details

Table 2 summarises architecture and training hyperparameters. Student stopping and selection use validation Cox loss and IBS, respectively (Algorithm 1). The split seed is 20261112 and the training master seed is 20261113. Seeds for each fold, stage and ensemble member are derived deterministically using SHA-256, with three independent teacher seeds per fold complement.

**Table 2.**
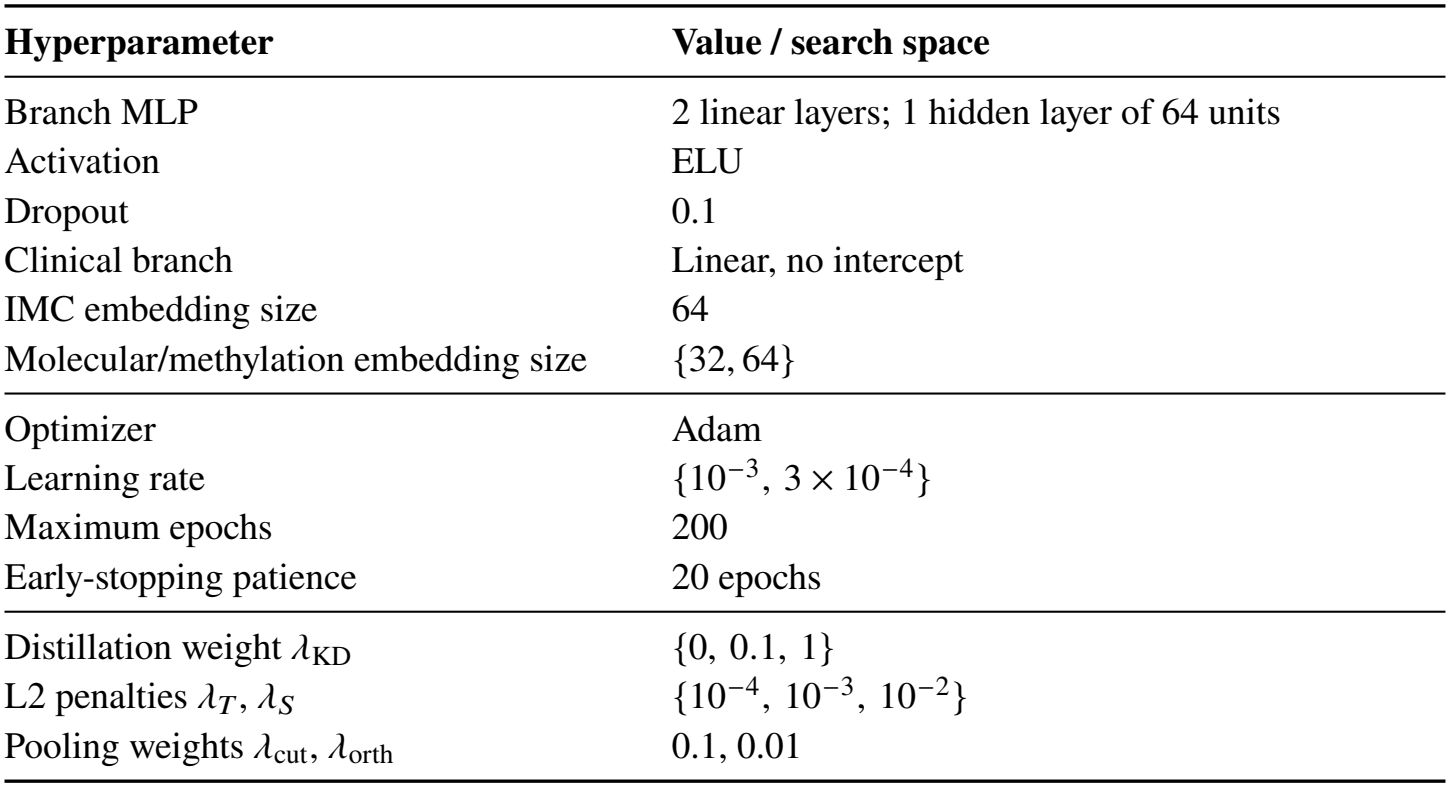
HEAL-Surv hyperparameters; braces denote search candidates.

### 4 Results and Discussion

### 4.1 DSS Prediction Performance

In METABRIC cross-validation (Table 3), the four-modality teacher achieved the highest mean concordance and the lowest mean Brier-based errors among the evaluated models. The modality ablations in Sect. 4.3 examine how each input contributes to these results. With architecture and inference inputs held fixed, distillation increased mean five-year Uno’s concordance from 0.658 to 0.704, close to DMGN’s 0.701. The distilled student used only molecular and clinical inputs for prediction. Its IBS rose from 0.071 to 0.074,and its five-year Brier score rose from 0.142 to 0.147.

**Table 3.** Within-cohort DSS performance (mean ±fold SD; 5-fold METABRIC, 3 ×5-fold Basel). Bold: best mean for each metric within each cohort.

| **Method** | **Harrell's C ↑** | **Uno's C ↑**<br>(5 years) | **IBS ↓**<br>(1-60 months) | **Brier score ↓**<br>(5 years) |
|---|---|---|---|---|
| **A. METABRIC** |  |  |  |  |
| *Reference methods* |  |  |  |  |
| CPH [10] | 0.599 ± 0.023 | 0.623 ± 0.039 | 0.071 ± 0.007 | 0.145 ± 0.017 |
| CPH (molecular + clinical) [10] | 0.657 ± 0.015 | 0.683 ± 0.020 | 0.073 ± 0.016 | 0.145 ± 0.030 |
| RSF [19] | 0.642 ± 0.026 | 0.672 ± 0.032 | 0.070 ± 0.007 | 0.141 ± 0.016 |
| DeepSurv [22] | 0.599 ± 0.023 | 0.637 ± 0.046 | 0.071 ± 0.007 | 0.145 ± 0.017 |
| DeepHit [23] | 0.560 ± 0.068 | 0.565 ± 0.050 | 0.073 ± 0.007 | 0.149 ± 0.016 |
| BroadSurv [31] | 0.578 ± 0.030 | 0.595 ± 0.043 | 0.072 ± 0.006 | 0.145 ± 0.012 |
| DAGSurv [27] | 0.508 ± 0.099 | 0.533 ± 0.076 | 0.073 ± 0.007 | 0.149 ± 0.014 |
| AttentionSurv [28] | 0.615 ± 0.029 | 0.664 ± 0.033 | 0.076 ± 0.013 | 0.154 ± 0.030 |
| DMGN [14] | 0.677 ± 0.027 | 0.701 ± 0.043 | 0.070 ± 0.009 | 0.144 ± 0.024 |
| *HEAL-Surv (this work)* |  |  |  |  |
| HEAL-Surv teacher | **0.703** ± **0.033** | **0.723** ± **0.045** | **0.060** ± **0.017** | **0.140** ± **0.024** |
| Student without distillation | 0.631 ± 0.042 | 0.658 ± 0.058 | 0.071 ± 0.016 | 0.142 ± 0.023 |
| HEAL-Surv student | 0.678 ± 0.031 | 0.704 ± 0.045 | 0.074 ± 0.019 | 0.147 ± 0.026 |
| **B. Basel** |  |  |  |  |
| *Reference methods* |  |  |  |  |
| CPH [10] | 0.601 ± 0.077 | 0.630 ± 0.064 | 0.063 ± 0.020 | 0.134 ± 0.022 |
| CPH (IMC protein features + clinical) [10] | 0.611 ± 0.044 | 0.638 ± 0.027 | 0.065 ± 0.013 | 0.138 ± 0.008 |
| RSF [19] | 0.607 ± 0.081 | 0.628 ± 0.099 | 0.064 ± 0.019 | 0.137 ± 0.023 |
| DeepSurv [22] | 0.602 ± 0.071 | 0.638 ± 0.083 | 0.062 ± 0.019 | 0.132 ± 0.023 |
| DeepHit [23] | 0.579 ± 0.072 | 0.600 ± 0.084 | 0.064 ± 0.020 | 0.137 ± 0.023 |
| BroadSurv [31] | 0.592 ± 0.067 | 0.607 ± 0.078 | 0.064 ± 0.019 | 0.134 ± 0.024 |
| DAGSurv [27] | 0.581 ± 0.071 | 0.599 ± 0.081 | 0.065 ± 0.019 | 0.138 ± 0.023 |
| AttentionSurv [28] | 0.615 ± 0.071 | 0.643 ± 0.076 | 0.074 ± 0.025 | 0.156 ± 0.037 |
| DMGN [14] | 0.430 ± 0.083 | 0.440 ± 0.083 | 0.067 ± 0.018 | 0.149 ± 0.026 |
| *HEAL-Surv (this work)* |  |  |  |  |
| HEAL-Surv spatial variant | **0.637** ± **0.059** | **0.655** ± **0.087** | **0.061** ± **0.013** | **0.131** ± **0.017** |

In repeated within-cohort cross-validation on Basel, the IMC-clinical spatial variant achieved the best mean results among the evaluated models on all four metrics, including a mean five-year Uno’s concordance of 0.655.

### 4.2 External Validation

We trained the shared-input models on METABRIC and evaluated them on Basel without refitting (Table 4). The HEAL-Surv spatial variant achieved the highest concordance, with a five-year Uno’s C of 0.660. Its probability errors were within the range of the clinical references. DeepHit had the lowest IBS, and AttentionSurv had the lowest five-year Brier score.

**Table 4.** METABRIC-to-Basel external validation (estimate ±patient-bootstrap SD). Bold: best estimate for each metric.

| **Method** | **Harrell's C ↑** | **Uno's C ↑**<br>(5 years) | **IBS ↓**<br>(1-60 months) | **Brier score ↓**<br>(5 years) |
|---|---|---|---|---|
| *Shared clinical inputs: age, ER, CT, HT* |  |  |  |  |
| CPH [10] | 0.614 ± 0.038 | 0.625 ± 0.043 | 0.064 ± 0.009 | 0.135 ± 0.015 |
| RSF [19] | 0.564 ± 0.043 | 0.567 ± 0.050 | 0.065 ± 0.008 | 0.142 ± 0.015 |
| DeepSurv [22] | 0.610 ± 0.037 | 0.619 ± 0.042 | 0.065 ± 0.009 | 0.136 ± 0.015 |
| DeepHit [23] | 0.590 ± 0.037 | 0.588 ± 0.043 | **0.063** ± **0.009** | 0.135 ± 0.015 |
| BroadSurv [31] | 0.601 ± 0.039 | 0.605 ± 0.046 | 0.064 ± 0.008 | 0.135 ± 0.014 |
| DAGSurv [27] | 0.606 ± 0.038 | 0.609 ± 0.044 | 0.065 ± 0.009 | 0.137 ± 0.015 |
| AttentionSurv [28] | 0.628 ± 0.038 | 0.648 ± 0.043 | 0.067 ± 0.008 | **0.131** ± **0.014** |
| *Shared clinical inputs and IMC* |  |  |  |  |
| DMGN (shared-input variant) [14] | 0.550 ± 0.048 | 0.536 ± 0.056 | 0.075 ± 0.011 | 0.153 ± 0.019 |
| HEAL-Surv spatial variant | **0.637** ± **0.042** | **0.660** ± **0.048** | 0.064 ± 0.009 | 0.136 ± 0.017 |

DMGN is the closest comparator by input modality because both spatial models use shared clinical variables and IMC. Most other references use clinical variables alone. HEAL-Surv achieved higher concordance and lower Brier-based errors than the shared-input DMGN variant. These results characterise the external performance of the IMC-clinical variant. The full teacher and the molecular-clinical student still require external validation.

### 4.3 Ablation Studies

Removing any modality reduced mean concordance and increased both Brier-based errors (Table 5). Removing the molecular branch produced the largest deterioration across the reported metrics, lowering mean five-year Uno’s C from 0.723 to 0.624. Removing IMC produced the smallest decrease in concordance, to 0.702, and increased both probability errors. Removing methylation or the clinical inputs, including ER, also worsened the reported means.

**Table 5.** METABRIC teacher modality ablation (mean ± fold SD, five folds).

| **Method** | **Harrell's C ↑** | **Uno's C ↑**<br>(5 years) | **IBS ↓**<br>(1-60 months) | **Brier score ↓**<br>(5 years) |
|---|---|---|---|---|
| All modalities | **0.703 ± 0.033** | **0.723 ± 0.045** | **0.060 ± 0.017** | **0.140 ± 0.024** |
| w/o clinical (incl. ER) | 0.664 ± 0.088 | 0.679 ± 0.103 | 0.075 ± 0.010 | 0.152 ± 0.022 |
| w/o molecular | 0.607 ± 0.042 | 0.624 ± 0.093 | 0.077 ± 0.010 | 0.160 ± 0.030 |
| w/o methylation | 0.665 ± 0.037 | 0.685 ± 0.101 | 0.076 ± 0.011 | 0.158 ± 0.030 |
| w/o IMC | 0.682 ± 0.043 | 0.702 ± 0.070 | 0.076 ± 0.009 | 0.155 ± 0.031 |

Stability-weighted distillation improved all four reported means relative to uniform distillation (Table 6A). Uniform distillation performed worse than training on observed outcomes alone. Relative to that outcome-only baseline, the stability-weighted student achieved higher concordance and had higher IBS and five-year Brier scores. Distillation from teachers with different input sets would help quantify how much IMC and methylation contribute to the gain in risk ranking.

**Table 6.** METABRIC component ablations (mean ± fold SD, five folds).

| **Configuration** | **Harrell’s C ↑** | **Uno’s C ↑**<br>(5 years) | **IBS ↓**<br>(1-60 months) | **Brier score ↓**<br>(5 years) |
|---|---|---|---|---|
| **A. Student distillation** |  |  |  |  |
| Without KD | 0.631 ± 0.042 | 0.658 ± 0.058 | 0.071 ± 0.016 | 0.142 ± 0.023 |
| Uniform KD | 0.560 ± 0.030 | 0.575 ± 0.028 | 0.077 ± 0.017 | 0.154 ± 0.031 |
| Stability-weighted KD | 0.678 ± 0.031 | 0.704 ± 0.045 | 0.074 ± 0.019 | 0.147 ± 0.026 |
| **B. Teacher graph encoder** |  |  |  |  |
| Non-hierarchical | 0.603 ± 0.073 | 0.618 ± 0.067 | 0.073 ± 0.017 | 0.148 ± 0.026 |
| Hierarchical | 0.703 ± 0.033 | 0.723 ± 0.045 | 0.060 ± 0.017 | 0.140 ± 0.024 |

The hierarchical teacher achieved higher mean concordance and lower mean Brier-based errors than the flat teacher (Table 6B), with mean five-year Uno’s C increasing from 0.618 to 0.723. Both concordance indices had smaller fold SDs. The comparison evaluates the hierarchical branch as a whole, including its pooled representations, coarse message passing, and pooling penalties.

### 4.4 Patient-Level Illustration

Figure 2 pairs predicted net DSS curves with IMC channels, niche labels, and local edge contributions for two held-out METABRIC patients selected by outcome. The CPH reference based on clinical variables and summary features assigned the two patients similar risks. HEAL-Surv estimated lower net DSS for MB-0288, who died of breast cancer, than for MB-2933, who died of another cause. The figure displays predictions through year 10 for illustration. The cohort-level evaluations use the primary five-year horizon.

**Fig 2.**
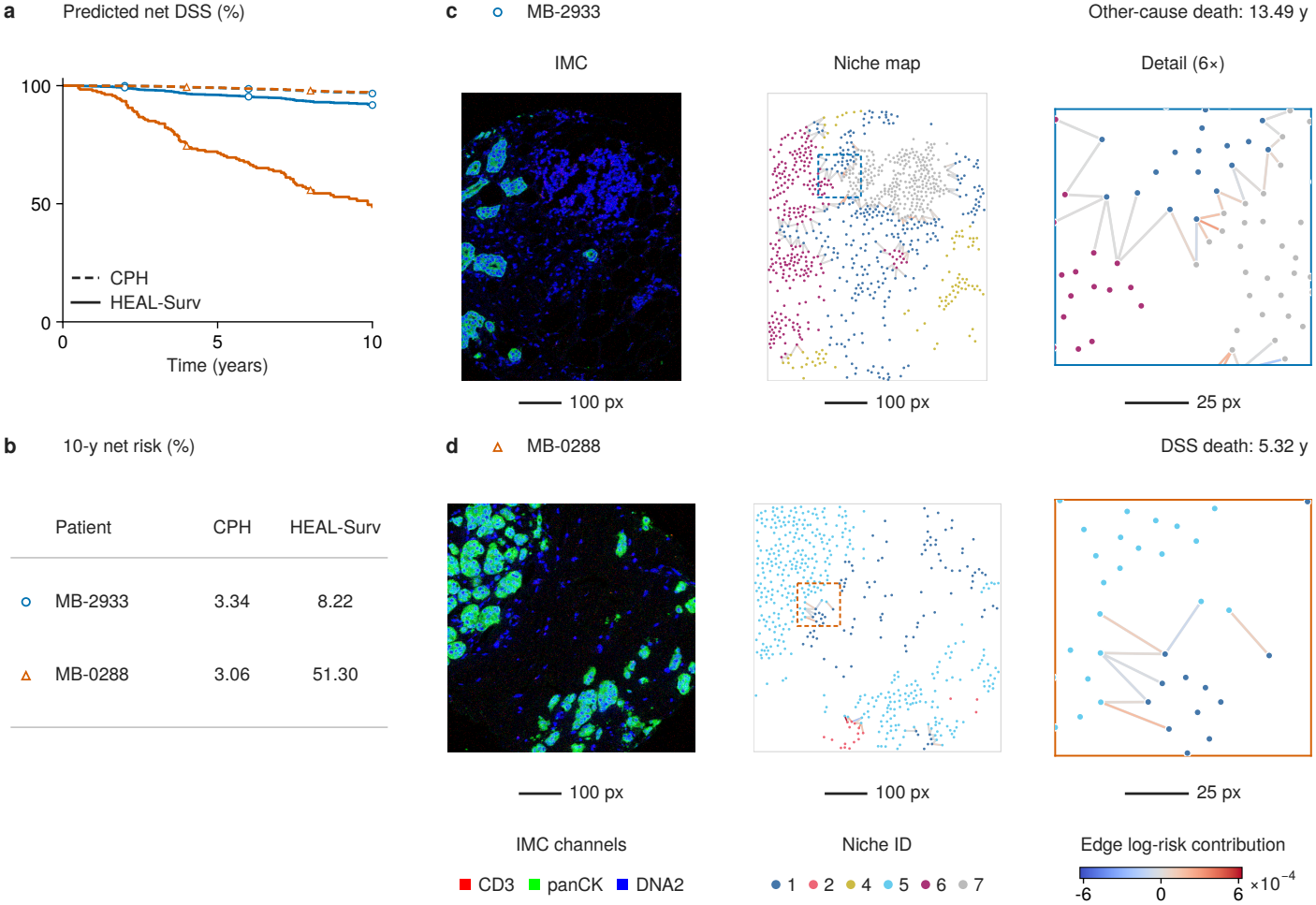
Ten-year net DSS predictions and tissue maps for two held-out METABRIC patients selected by outcome. MB-0288 died of breast cancer 5.32 years after diagnosis, and MB-2933 died of another cause 13.49 years after diagnosis.

### 4.5 Spatial Interpretation

Using the frozen IMC-clinical model, we examined how tissue arrangement relates to predicted log-risk in Basel. We quantified immune-epithelial mixing by comparing observed edge counts with their expectation under permutation of cell-type labels. Let *O*_*ir*_ denote the observed number of immune-epithelial edges in core *r* of patient *i*, and let ℰ_*ir*_ denote its undirected edges excluding self-loops. With 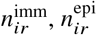, and *n*_*ir*_ denoting immune, epithelial, and total cell counts, respectively, the expected edge count and patient-level mixing score are

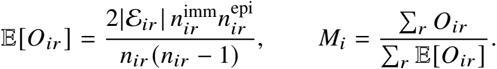

The patient-level score *M*_*i*_ divides the total observed edge count across the patient’s cores by the total expected count.

Figure 3(a,b) shows the largest cores from patients nearest the lower and upper quartiles of *M*_*i*_. Selection was based on *M*_*i*_ alone. The higher-mixing example has fewer observed immune-epithelial edges and a lower expected count, illustrating how the expected count affects the comparison. Cells are shown as centroid-based Voronoi regions clipped at 7.5 *µ*m. We estimated the partial Spearman correlation between *M*_*i*_ and IMC log-risk 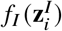 , adjusting for immune and epithelial cell fractions and ER status. Ranks and adjustment were recomputed in 2,000 patient-level bootstrap samples. The adjusted correlation was 0.04 (95% CI ™0.09 to 0.16), indicating a weak estimated association with an interval that includes zero (Fig. 3c).

**Fig 3.**
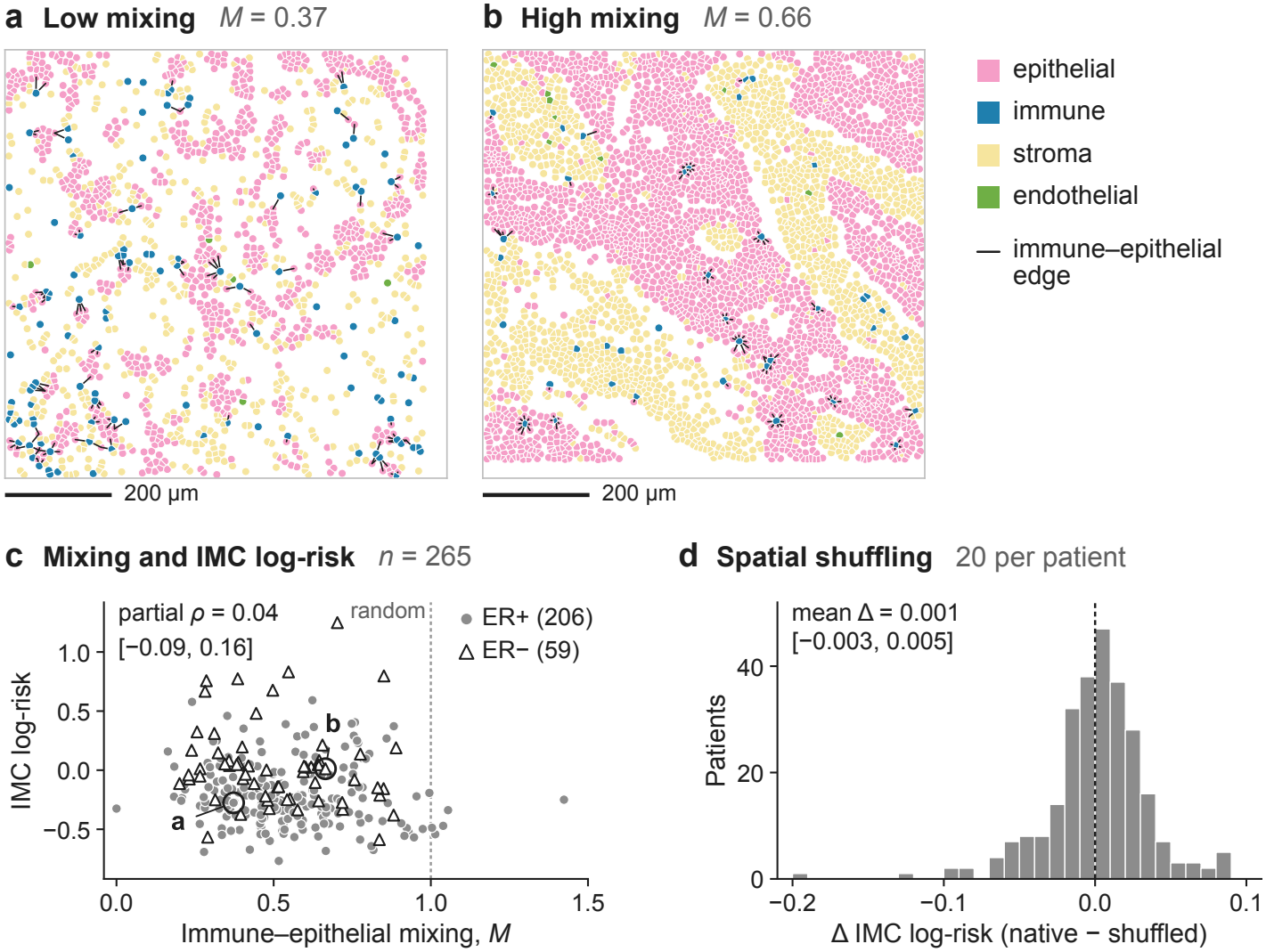
Spatial analysis of the frozen IMC-clinical model in Basel: (a,b) relatively low/high mixing;(c) mixing versus IMC log-risk; (d) native minus mean shuffled log-risk. The example cores contain 104 and 95 observed immune-epithelial edges, versus expected counts of 279 and 143, respectively. Rings: example patients; brackets: 95% patient-bootstrap intervals.

To probe spatial sensitivity, we jointly shuffled protein vectors and cell-type labels across fixed positions, preserving cellular profiles and composition. Across 20 repetitions per core, the mean absolute change in IMC log-risk was 0.0235 (95% CI 0.0209-0.0266). Figure 3(d) shows the signed difference between native and mean shuffled log-risk. The shuffling analysis probes sensitivity to cellular arrangement, while the encoder ablation evaluates the predictive contribution of hierarchical encoding.

## 5 Conclusion

We developed HEAL-Surv, a teacher-student framework that combines hierarchical IMC graph learning with multimodal distillation for five-year net DSS prediction. In METABRIC, the four-modality teacher achieved the strongest mean performance among the evaluated models. Stability-weighted distillation increased the matched student’s mean five-year Uno’s concordance from 0.658 to 0.704 using only molecular and clinical inputs at inference. The distilled student had higher Brier-based errors. Ablations supported the complete teacher and its hierarchical IMC encoder. In a separate external evaluation on Basel, the IMC-clinical variant achieved a five-year Uno’s concordance of 0.660. External validation and direct calibration assessment of the molecular-clinical student remain priorities.

## Data Availability

All source data analysed in this study are from previously published cohorts and are openly available. The Basel imaging mass cytometry cohort (Jackson et al., Nature 2020) is available on Zenodo (https://doi.org/10.5281/zenodo.3518284). The METABRIC imaging mass cytometry data are available on Zenodo (https://doi.org/10.5281/zenodo.5850952). The METABRIC clinical, molecular and DNA methylation data are available as described in the data availability statements of the original publications cited in the manuscript (Ali et al. 2020; Batra et al. 2021; Curtis et al. 2012; Pereira et al. 2016), and records were linked through the released METABRIC patient identifiers. No new data were generated.

## Acknowledgments

The authors acknowledge the support of the Hanoi Medical University-Australia Young Researchers Seed Fund.

